# Exploring Novel Biomarkers for Early Detection of Osteoporosis

**DOI:** 10.1101/2025.05.30.25328641

**Authors:** Filipe Ricardo Carvalho, Virna Ferreira Queiroz, Paulo Jorge Gavaia

**Affiliations:** Faculty of Medicine and Biomedical Sciences, University of Algarve, Faro, Portugal; Centre of Marine Sciences (CCMAR/CIMAR LA), University of Algarve, Faro, Portugal Universidade do Algarve - Campus de Gambelas, 8005-139 Faro, Portugal

**Author notes:** Corresponding author: Filipe Ricardo Carvalho.

**Keywords:** Osteoporosis, Machine Learning, Bone Mineral Density, Biomarkers, Magnesium

## Abstract

**Purpose/Introduction:** Osteoporosis is characterized by diminished BMD and deteriorated bone microstructure, significantly increasing fracture susceptibility. This study leverages machine learning for identifying key predictors of osteoporosis, potentially improving non-invasive screening tools.

**Methods:** We utilized the “Bone Mineral Density” dataset from Harvard University, categorizing variables into demographics, bone loss prevention treatments, and lifestyle/associated pathologies. A Boolean variable “OP” was defined to indicate presence of osteoporosis. Data preprocessing included imputation for missing values and normalization. Statistical analyses such as Welch’s T-test and multiple linear regression were conducted to identify significant markers. Additionally, unsupervised learning techniques, like Louvain clustering, were used to uncover natural data patterns, while supervised learning models aimed to predict osteoporosis.

**Results:** The study identified significant markers associated with osteoporosis, including magnesium levels, high- and low-density lipoproteins, chronic obstructive pulmonary disease, and chronic kidney disease. Multiple linear regression analysis revealed robust associations between osteoporosis and biomarkers such as ALT, BUN, CREA, LDL-C, and Mg, even after controlling for age, height, and weight. Among supervised learning models, Random Forest and Gradient Boosting algorithms showed the highest accuracy in predicting osteoporosis, with Random Forest identified as the preferred model.

**Conclusions:** In our work, it was possible to identify the main biochemical parameters and clinical histories that are potential predictors of osteoporosis. The findings suggest that integrating machine learning techniques with clinical data can enhance early detection and intervention strategies, ultimately improving patient outcomes. Further refinement is recommended to validate these results across diverse populations and improve predictive models for broader application.

## Introduction

Osteoporosis (OP) is a progressive skeletal disorder characterized by diminished bone mineral density (BMD) and deterioration of bone microstructure (1,2). This condition significantly increases bone fragility and susceptibility to fractures (3). While reduced BMD is a key indicator of fracture risk, other factors such as bone geometry, microarchitecture, and size also contribute to overall bone strength and resilience against trauma (2,4).

Often referred to as a “silent disease,” OP typically remains undetected until a fracture occurs, requiring imaging and radiologic tests for diagnosis.

Fragility fractures, the most evident clinical manifestation of OP, commonly occur in the hip, spine, and distal radius (5–7). The relationship between fracture risk and BMD varies between men and women, influenced by age-related differences in bone weakness progression (2,8,9).

The condition is classified into two main categories: primary OP, associated with age and decreased sex hormones (particularly estrogen), and secondary OP, linked to other pathologies or medical treatments (10).

Primary OP disproportionately affects women due to their naturally lower bone density and the impact of menopause. However, estrogen deficiency also plays a role in male OP, as bone cells in both sexes possess estrogen receptors. Men with OP are more likely to experience secondary forms of the disease, often due to underlying pathologies or medication-induced bone loss (5,11).

The elderly population, which is rapidly growing worldwide, faces the highest risk of OP. This demographic shift presents increasing social and economic challenges related to the disease. Age-related decline in bone mass and increased fracture risk are compounded by other factors such as low body mass index, alcohol and tobacco consumption, history of falls, previous fractures, family history of OP, and certain chronic and inflammatory diseases (12–14).

Understanding the complex interplay of factors contributing to OP is crucial for developing effective diagnostic, preventive, and treatment strategies (15,16). As the global population ages, the need for accurate, accessible, and non-invasive screening tools becomes increasingly urgent.

Artificial intelligence (AI) and machine learning (ML) have emerged as promising tools in addressing this urgent need, offering potential solutions for early detection and risk assessment of osteoporosis. Recent studies have explored various AI-driven approaches, including deep learning algorithms for analyzing bone densitometry images (17), convolutional neural networks for interpreting radiographs, and ensemble learning methods for integrating multiple risk factors (18–20). These AI tools have shown promising results in improving the accuracy of fracture risk prediction and osteoporosis diagnosis. For instance, some researchers have developed ML models that can predict BMD from routine CT scans, potentially reducing the need for specialized DXA scans (21). Others have utilized natural language processing to extract relevant information from electronic health records, enhancing the identification of high-risk individuals (22). Additionally, AI-powered clinical decision support systems have been proposed to assist healthcare providers in making more informed treatment decisions based on individual patient data and established clinical guidelines (15). As these technologies continue to evolve, they hold the potential to revolutionize osteoporosis screening and management, enabling more personalized and proactive approaches to bone health.

## Purpose

This study aims to explore the potential of machine learning techniques in identifying key biochemical and clinical markers associated with OP, potentially leading to improved early detection and intervention strategies.

## Methods

The data used in this study was from the “Bone mineral density” dataset (23) obtained from the Harvard Dataverse platform, developed by the Institute for Quantitative Social Science (IQSS) at Harvard University. The dataset comprises comprehensive clinical information, dual-energy X-ray absorptiometry (DXA) measurements, and various variables crucial for studying bone health, OP, and fracture risk.

Variables were categorized into three main groups (Table 1):

1. Demographics & Analytics: Personal characteristics and medical biomarkers.
2. Bone Loss Prevention Treatments: Interventions aimed at maintaining bone health.
3. Lifestyle/Associated Pathologies: Factors and conditions potentially correlated with OP (Figure 1).

**Table 1.**
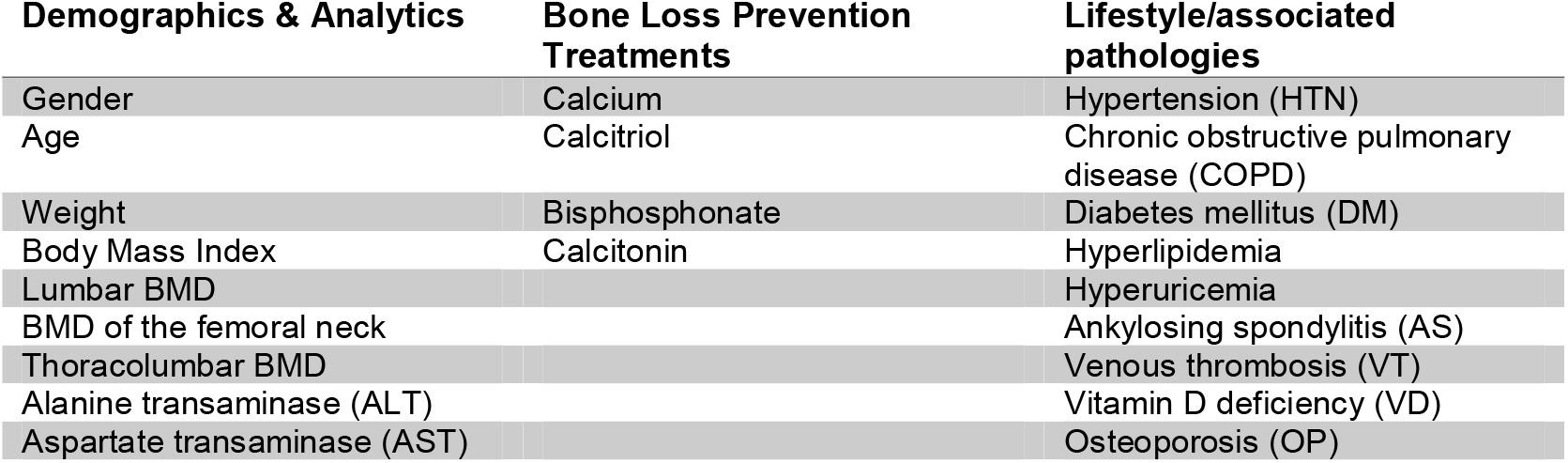
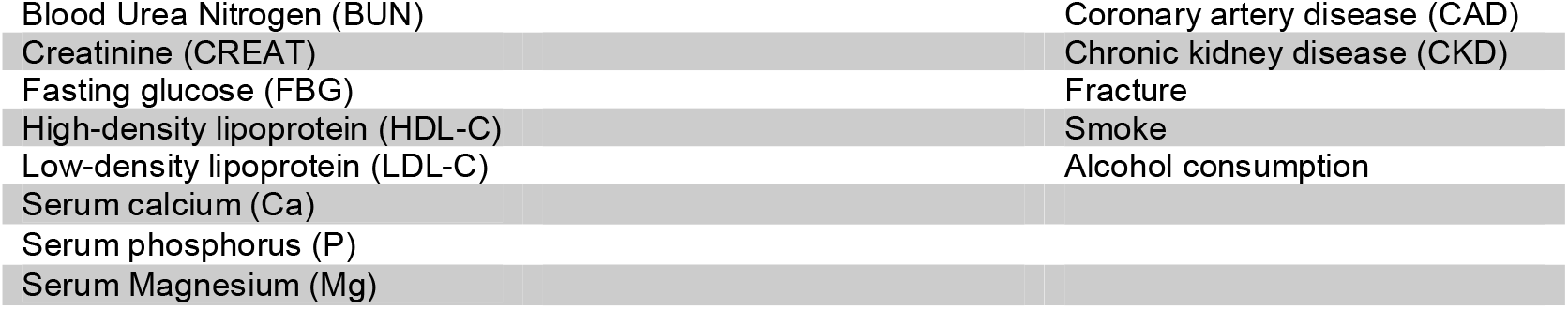
Organization into database groups *Bone mineral density*.

**Figure 1.**
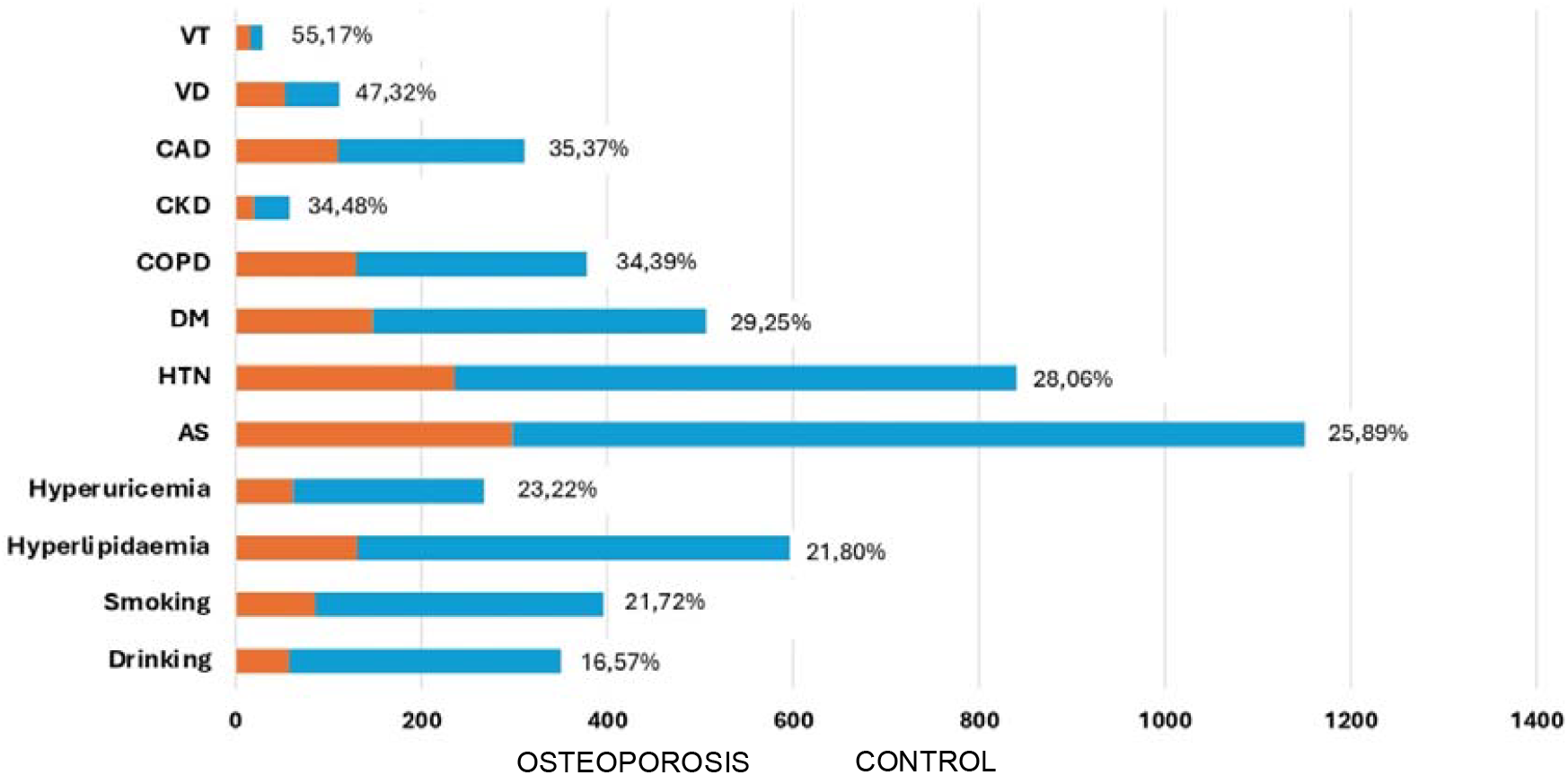
Comparison of pathologies between the Osteoporosis (OP) and Control groups. VT: venous thrombosis; RV: vitamin D deficiency; DKA: coronary artery disease; CKD: chronic kidney disease; COPD: chronic obstructive pulmonary disease; DM: diabetes mellitus; HTN: hypertension; AS: ankylosing spondylitis.

### 1. Defining the Target Variable

Due to the absence of metadata defining OP in the original dataset, we created a Boolean variable “OP” (Figure 2). This variable was set to True if a participant met any of the following criteria:

**Figure 2.**
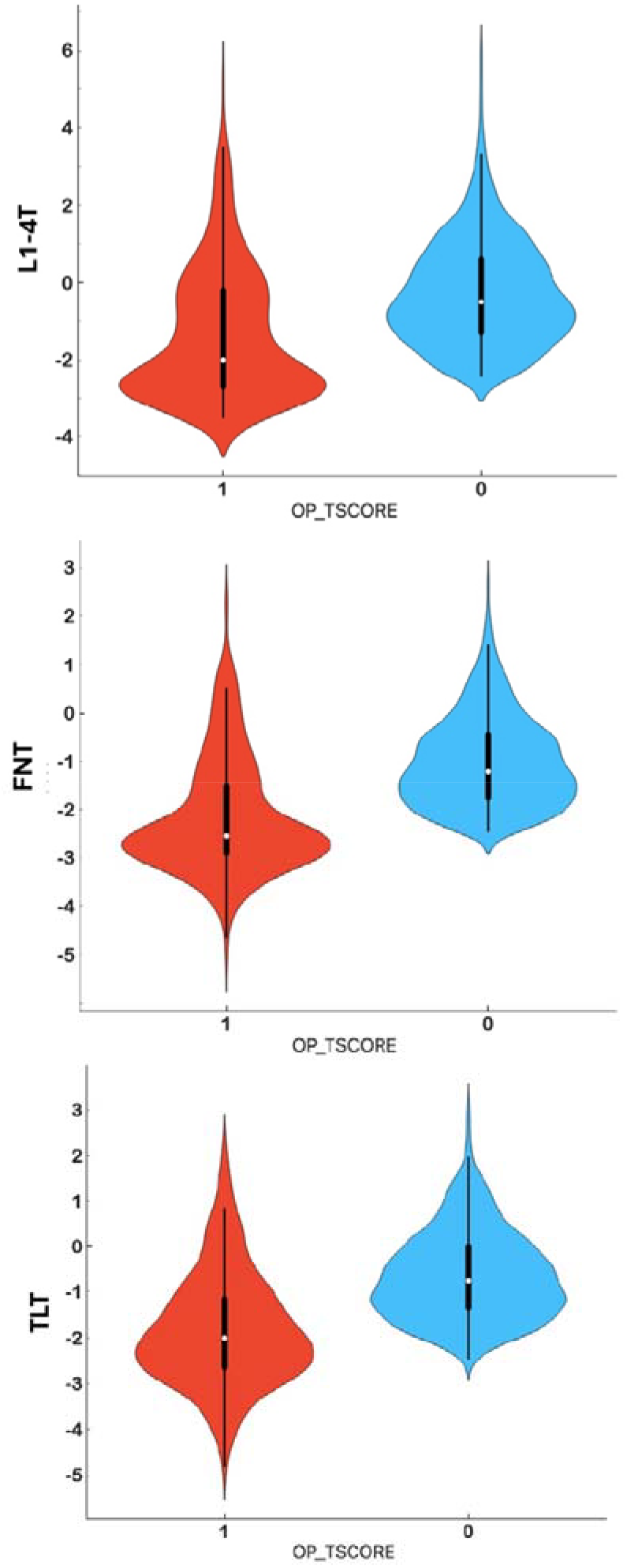
Violin plot comparing T-score values between individuals with osteoporosis (OP) and a control group across three key skeletal regions: the thoracolumbar region (TLT), femoral neck (FNT), and lumbar vertebrae L1 to L4 (L1-4T) The plot’s shape illustrates the distribution of T-score differences, with wider sections indicating higher frequency of data points at those values. TLT in the control group averaged −0.64 (SD 0.97), while in the OP group it was −1.82 (SD 1.22). FNT showed −1.02 (SD 0.93) for controls and −2.17 (SD 1.17) for the OP group. L1-4T averaged −0.27 (SD 1.36) in controls and −1.40 (SD 1.68) in the OP group. These statistics, visualized through the violin plot, highlight the consistently lower bone density and greater variability in T-scores among osteoporosis patients across all measured skeletal regions.

- T-score ≤ −2.5 in at least one of the tested locations: femoral neck (TNF), thoracolumbar region (TLT), or L1-L4 (L1.4T).
- History of fractures, this definition aligns with established guidelines (2).

### 2. Data pre-processing

#### 2.1. Missing data filling and Data normalization

Imputation Missing values were imputed using the mean of each variable. For certain analyses, data were normalized to a 0-1 scale to standardize values across variables, facilitating comparisons. This process transforms the minimum value to 0 and the maximum to 1, with intermediate values adjusted proportionally.

#### 2.2. Statistical Analysis with Welch’s T-Test

Welch’s t-test was employed to compare means between the OP and control groups for each variable. This test is suitable for samples with potentially unequal variances or sizes. Two-tailed P-values were calculated, with P < 0.05 considered statistically significant. The analysis was performed on the entire sample and separately for men and women to identify gender-specific differences (Table 2).

**Table 2.**
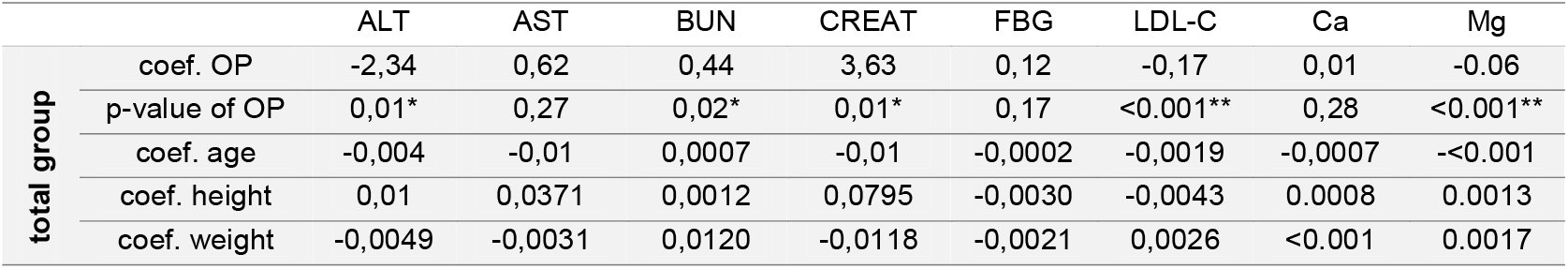
Results of the coefficients and p-values of the multiple linear regression analysis. p<0.05;**p<0.001 Coef.: Coefficient.

### 3. Correlation analysis

A correlation matrix was generated and visualized as a heatmap to explore relationships between variables, with a focus on their associations with OP and BMD. This analysis intended to establish patterns and identify key correlations among the dataset parameters (supplementary material Figure 5).

### 4. Comparison Between Groups and Control of Covariates

To investigate the relationship between OP (represented by the independent variable OP) and various biochemical variables (dependent variables), while accounting for covariates such as age, height, and weight, we performed multiple linear regression analyses. This approach allowed us to control for the effects of covariates and understand how their variances affect the generated models.

We selected the following dependent variables for analysis: alanine aminotransferase (ALT), aspartate aminotransferase (AST), blood urea nitrogen (BUN), blood creatinine (CREA), uric acid (URIC), fasting blood glucose (FBG), high-density cholesterol (HDL-C), low-density cholesterol (LDL-C), serum levels of calcium (Ca), phosphorus (P), and magnesium (Mg). The independent variable was OP, with age, height, and weight as covariates.

This method enabled us to apply a T-test and determine whether the coefficients significantly impact the prediction of each dependent variable. We applied this analysis to the total sample, as well as separately female and male groups, to identify any gender-specific differences.

### 5. Prevalence of OP in Different Pathological Conditions

Recognizing that secondary OP can occur in relation to other pathologies and lifestyle factors, we analyzed the prevalence of OP among subjects with various comorbidities. We organized the incidence of OP in relation to the presence of diseases or lifestyles considered to be risk factors in both the OP and control groups (Figure 1).

### 6. Unsupervised Learning Analytics

We employed unsupervised machine learning techniques to identify natural patterns in the data, particularly focusing on variables correlated with the T-score associated with OP. The Louvain Clustering method was chosen for this analysis.

First, we normalized the data and applied Principal Component Analysis (PCA) to reduce dimensionality. PCA identifies the directions of maximum variance, reducing redundancy and noise in the data. This allowed us to select the main components for the elaboration of the RadViz multivariate data visualization algorithm.

We then created a visual representation of connections between objects using k-nearest neighbors (KNN) with Euclidean distance metric and 17 neighbors. The Louvain community detection algorithm was applied to this graph to optimize modularity and find highly interconnected node communities into clusters based on their natural similarities.

### 7. Development of a Predictive OP Model through Supervised Learning

For our supervised learning approach, we developed a predictive model for OP. To ensure model efficiency and avoid redundancy, we excluded variables related to bone density values and potential treatments (calcium, calcitriol, and bisphosphonates).

We divided the data into a training set (75% of the data) and a test set (25%). We then tested eight different classification algorithms to determine which would perform best in predicting the True classes of OP: Gradient Boosting, Random Forest, Decision Tree, Logistic Regression, AdaBoost, Neural Network, Support Vector Machine (SVM), Naive Bayes.

To evaluate the results of the developed models, we used the following metrics: Area under the ROC curve (AUC), Classification Accuracy (CA), F1 score (precision-weighted harmonic mean and recall), Precision (proportion of true positives among instances classified as positive), Recall (ratio of true positives among all positive instances) and Matthews Correlation Coefficient (MCC). Finally, we identified the importance of features that most strongly influenced the results of the chosen algorithm for predicting the diagnosis.

### 8. Data Analysis Tools

We used Orange Data Mining software (version 3.36.2) (24) for all data analyses. The specific plugins employed include Orange Data Table: For importing, cleaning, and preparing the raw data, Louvain Clustering: For unsupervised learning analysis, Correlations and Heat Map: For correlation analysis and visualization, Modeling Plugins: For implementing supervised learning algorithms, Test and Score: For evaluating algorithmic performance on test data. Feature Importance: For analyzing variable influence on model accuracy

## Results

### 1. Pathologies, lifestyle and osteoporosis

The prevalence of OP in relation to various pathologies and lifestyle factors within our study population is present in Figure 1. Among the variables examined, venous thrombosis showed the highest co-occurrence with OP, with 55.17% of individuals with this condition. Vitamin D deficiency was the second most prevalent condition, with 47.32% followed by coronary artery disease with 35.37%, while OP cases were observed in chronic kidney disease and chronic obstructive pulmonary disease patients in 34.84% and 34.39%, respectively the distribution of T-scores in relation to control and OP groups. A Violin Plot was constructed to visualize this distribution of the density curves of the T-scores for L1-4T, TNF, TLT regions, allowing for a comparison of data distribution between the OP and non-OP groups (Figure 2).

### 2. Biochemical Data Results

To analyze the biochemical parameters and their behaviors between OP and control groups, we used Welch’s t-test and multiple linear regression to assess the impact of covariates. Significant differences were found for ALT, were the OP group showed mean value of 22.86 with a standard deviation (SD) of 13.91, compared to 23.77 (SD = 17.41) in the control group. Significant differences were also found in BUN OP averaging 5.52 (SD = 1.85) and the control group at 5.65 (SD = 3.67). In CREAT with a mean of 74.49 (SD = 26.69) in OP versus 73.81 (SD = 25.36) for the controls. Notably, LDL-C levels average 2.53 (SD = 0.91) against the control’s 2.62 (SD = 0.89). Magnesium (Mg) presented the most notable variation, with the OP group averaging lower at 0.83 (SD = 0.11) compared to 0.88 (SD = 0.09) in the control group (Figure 3).

**Figure 3.**
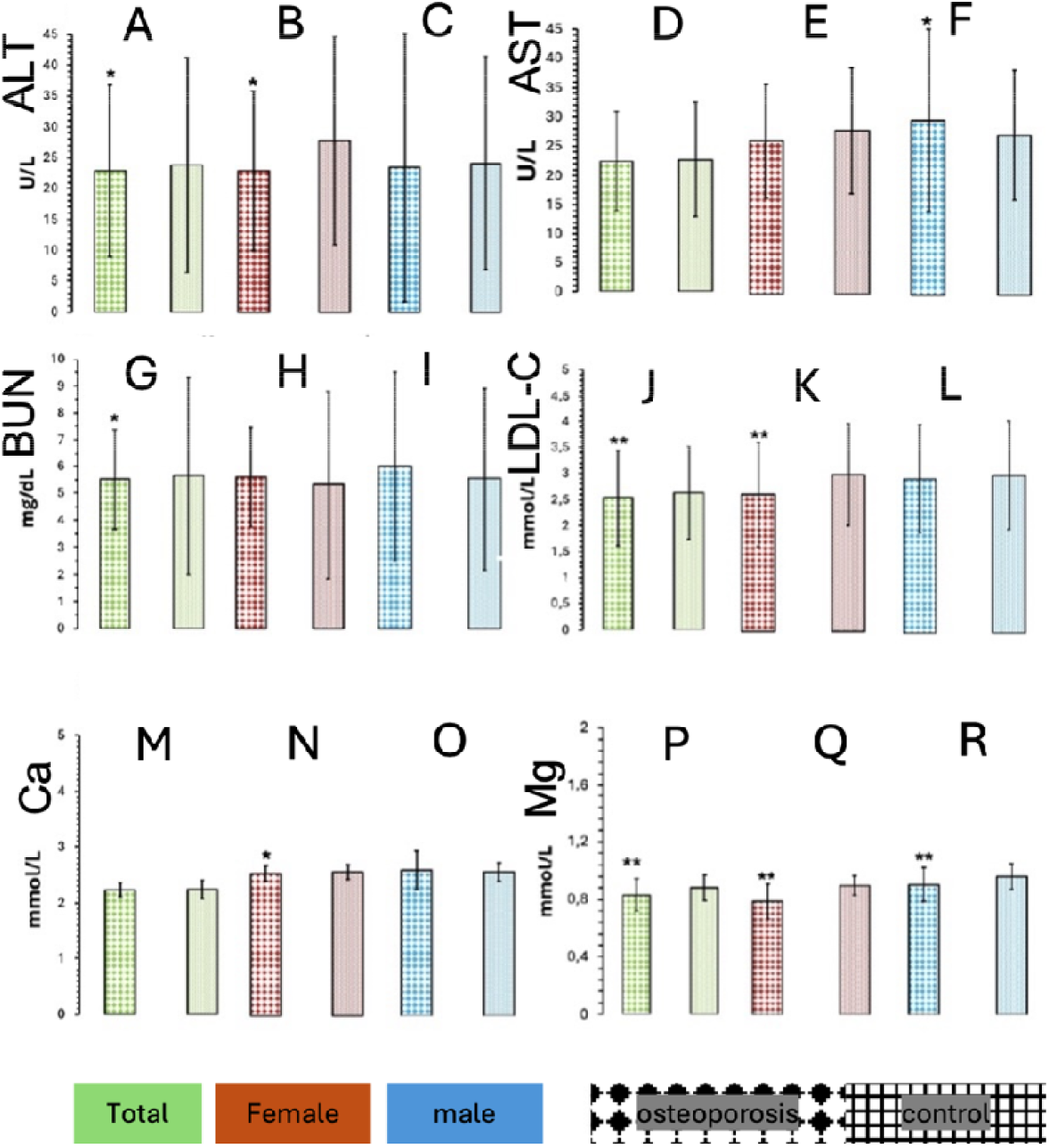
Mean concentration values of significant biomarkers. A, B, C: Alanine aminotransferase (ALT) values. For the Total and Female groups, significant differences were found for *p<0.05. D, E, F: Aspartate aminotransferase (AST) values. For the male group, significant differences were found for *p<0.05. G, H, I: Nitrogen urea (BUN) values. For the Total groups, significant differences were found for *p<0.05. J, K, L: Low-density lipoprotein (LDL-C) values. For the Total and Female groups, significant differences were found for **p<0.001. M, N, O: Serum calcium (Ca) values. For the female group, significant differences were found for *p<0.05. P, Q, R: Serum magnesium (Mg) values. For all groups – Total, Female and Male – significant differences were found for **p<0.001.

The coefficients for age across all biomarkers were relatively small and did not indicate significant associations. The height and weight coefficients were not significant for most biomarkers, except for Mg, where both height and weight had significant positive influences. The significant relationships between OP and the biomarkers ALT, BUN, CREAT, and LDL-C remained robust even after controlling for age, height, and weight, indicating that the associations of these biomarkers with OP are independent of these potential confounding variables. However, for Mg, the significant positive influences of height and weight suggest that its association with OP is influenced by these covariates (supplementary material, Table 3).

### 3. Correlation of variables

Correlation analysis (supplementary material Figure 5), illustrates the correlations between various variables, emphasizing their associations with bone parameters. OP was inversely correlated with bone mineral density (BMD) and T-score. Treatments such as calcium, calcitriol, bisphosphonates, and calcitonin also show inverse correlations with BMD and T-scores of the femoral neck and thoracolumbar regions. These findings are consistent with our expectations, given the associations of these variables with bone health.

Mg exhibited significant correlations: a strong negative correlation with OP and direct correlations with BMD and T-scores of the lumbar region, indicating its potential as an evaluation parameter. Additionally, a subtle positive association was observed between alanine aminotransferase (ALT) and aspartate aminotransferase (AST) with bone health; ALT correlates with the femoral neck, femoral neck T-score, thoracolumbar region, and thoracolumbar T-score.

Conversely, uric acid (URIC) and high-density lipoprotein cholesterol (HDL-C) displayed opposite behaviors. URIC showed positive correlations with bone health parameters, particularly the femoral neck and thoracolumbar region, while HDL-C was inversely correlated with bone components.

### 4. Analysis of unsupervised learning outcomes

Louvain clustering identified four clusters from the data, as visualized in the Radviz plot in (supplementary material Figure 6). This analysis aimed to explore variable behaviors based on similarities, without targeted algorithmic interventions.

Cluster 1 (C1) includes OP, HDL-C, LDL-C, BUN, TLT, FNT, L1-4T, TL, FN, P, Mg, and Gender. Cluster 2 (C2) comprises BMI, Age, Weight, URIC, CREAT, Height, AST, and ALT. Cluster 3 (C3) contains L1-4, while Cluster 4 (C4) consists of FBG.

The inclusion of OP in C1 is particularly significant, revealing a hidden pattern among these variables. To ensure unbiased clustering of biochemical parameters, data on diseases or treatments were excluded.

### 5. Analysis of the different supervised learning models

Among the tested algorithms (supplementary material, Table 4), Gradient Boosting yielded the highest accuracy, closely followed by Random Forest. Both demonstrated strong performance in correctly identifying OP diagnoses, while other algorithms showed decreasing predictive accuracy, with Naive Bayes performing poorest. Confusion matrices (supplementary material Figure 7) for Gradient Boosting and Random Forest models, based on the 25% test data, revealed their respective performances: Gradient Boosting: 328 correct diagnoses (39 true positives for OP), 56 misclassifications (50 false negatives, 6 false positives). Random Forest: More false positives (14) but fewer false negatives (47), Higher true positives (42) compared to Gradient Boosting Due to Random Forest’s superior ability to identify OP, it was led to its selected as the preferred predictive model. Feature importance analysis (Figure 4) highlighted the most influential characteristics for the Random Forest model, which were Mg, COPD, history of fractures, height, and CAD.

**Figure 4.**
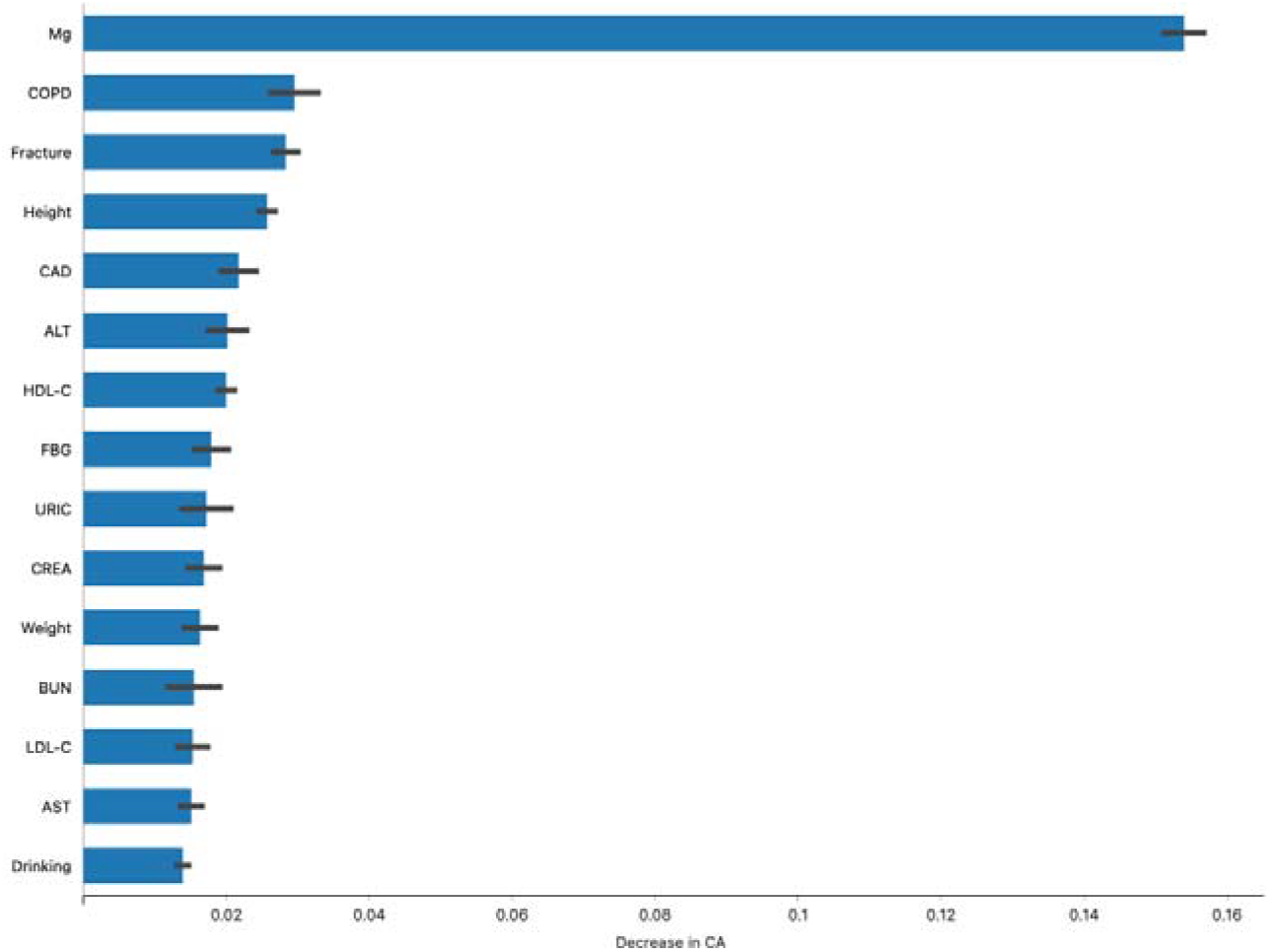
Feature Importance in the Random Forest Model. Figure illustrates the relative importance of various features in the Random Forest model for predicting osteoporosis (OP) diagnoses. The analysis identifies Magnesium (Mg), Chronic Obstructive Pulmonary Disease (COPD), history of fractures, height, and Coronary Artery Disease (CAD) as the most influential variables. These features significantly contribute to the model’s predictive accuracy, highlighting their critical roles in the assessment and diagnosis of osteoporosis.

## Conclusion

Our study aimed to identify key biochemical and clinical markers associated with OP using machine learning techniques. The results highlight several important findings and areas for further investigation.

Magnesium emerged as a significant predictor of bone health, showing strong correlations with BMD and OP. This aligns with previous research indicating magnesium’s crucial role in bone development and matrix formation (25,26). The highly significant values found between Mg and OP, along with the exceptionally strong correlation between Mg and especially L1-4 BMD, warrant further in-depth analysis. Despite the need for additional research, Mg demonstrates robust potential as a predictor of BMD.

However, it is critical to note that in our multiple linear regression analysis, the significance of magnesium’s association with OP was influenced by covariates such as age, height, and weight. These factors contribute to bone health and could potentially confound the direct impact of Mg. Age, in particular, is a well-known determinant of bone density changes (27–29), while height (30,31) and weight (32,33) influence biomechanical stress on the skeleton (34–36), affecting bone remodeling processes. The interplay of these covariates suggests that while Mg remains an important factor, its independent predictive power for bone health might be overestimated if these variables are not adequately controlled.

Furthermore, the clinical diagnosis of Mg deficiency remains challenging, as serum levels may not accurately reflect total body Mg status (37). This underscores the importance of developing more accurate methods to assess Mg levels in the context of bone health evaluation. Therefore, while magnesium’s role in bone health is significant(38), its assessment and the influence of other physiological factors must be carefully considered in both clinical and research settings to fully understand its impact.

Lipid profiles, particularly HDL-C and LDL-C, demonstrated noteworthy associations with bone health. Low HDL-C levels have been linked to impaired osteoblast differentiation and function (39) high LDL-C levels may promote osteoclastogenesis (40). These findings suggest a potential connection between lipid metabolism and bone health, which warrants further investigation.

Liver enzymes, especially ALT, showed relevance in our predictive model. This association may be explained by the link between chronic liver diseases and OP, known as hepatic osteodystrophy (41). However, the specific mechanisms underlying this relationship require further elucidation.

Uric acid demonstrated a positive correlation with bone health parameters, supporting previous findings of its potential protective effect on BMD in the elderly (42). However, the relationship between uric acid and bone health appears complex and may be influenced by gender and age.

The prevalence of OP in individuals with comorbidities such as chronic obstructive pulmonary disease (COPD) and chronic kidney disease (CKD) aligns with existing literature. These conditions share risk factors with OP and may involve treatments that affect bone metabolism (43,44).

This study demonstrates the potential of machine learning techniques in identifying key biochemical and clinical markers associated with OP. Magnesium, lipid profiles, liver enzymes, and uric acid emerged as significant predictors, alongside established risk factors such as age and comorbidities.

While our findings provide valuable insights, several limitations must be acknowledged. The cross-sectional nature of the data limits causal inferences, and the generalizability of the results to diverse populations requires further investigation. Additionally, the predictive model, while promising, needs further refinement to improve its accuracy and clinical utility. Future research should focus on longitudinal studies to establish causal relationships between identified markers and OP development. Refinement of machine learning models to enhance predictive accuracy and reduce false negatives. Validation of findings in diverse populations to ensure broad applicability. This study provides a foundation for developing more comprehensive and accurate screening tools for OP. By integrating biochemical markers with clinical risk factors, we may improve early detection and intervention strategies, ultimately reducing the burden of OP-related fractures and improving patient outcomes.

## Data Availability

All data produced in the present work are contained in the manuscript

## Acknowledgements

This study received Portuguese national funds from FCT - Foundation for Science and Technology through projects UIDB/04326/2020 (DOI:10.54499/UIDB/04326/2020), UIDP/04326/2020 (DOI:10.54499/UIDP/04326/2020) and LA/P/0101/2020 (DOI:10.54499/LA/P/0101/2020).

